# Characteristics and risk factors for COVID-19 diagnosis and adverse outcomes in Mexico: an analysis of 89,756 laboratory–confirmed COVID-19 cases

**DOI:** 10.1101/2020.06.04.20122481

**Authors:** Theodoros V Giannouchos, Roberto A Sussman, José Manuel Mier Odriozola, Konstantinos Poulas, Konstantinos Farsalinos

## Abstract

**Background:** There is insufficient information about risk factors for COVID-19 diagnosis and adverse outcomes from low and middle-income countries (LMICs).

**Objectives:** We estimated the association between patients’ characteristics and COVID-19 diagnosis, hospitalization and adverse outcome in Mexico.

**Methods:** This retrospective case series used a publicly available nation-level dataset released on May 31, 2020 by the Mexican Ministry of Health, with patients classified as suspected cases of viral respiratory disease. Patients with COVID-19 were laboratory-confirmed. Their profile was stratified by COVID-19 diagnosis or not. Differences among COVID-19 patients based on two separate clinical endpoints, hospitalization and adverse outcome, were examined. Multivariate logistic regressions examined the associations between patient characteristics and hospitalization and adverse outcome.

**Results:** Overall, 236,439 patients were included, with 89,756 (38.0%) being diagnosed with COVID-19. COVID-19 patients were disproportionately older, males and with increased prevalence of one or more comorbidities, particularly diabetes, obesity, and hypertension. Age, male gender, diabetes, obesity and having one or more comorbidities were independently associated with laboratory-confirmed COVID-19. Current smokers were 23% less likely to be diagnosed with COVID-19 compared to non-smokers. Of all COVID-19 patients, 34.8% were hospitalized and 13.0% experienced an adverse outcome. Male gender, older age, having one or more comorbidities, and chronic renal disease, diabetes, obesity, COPD, immunosuppression and hypertension were associated with hospitalization and adverse outcome. Current smoking was not associated with adverse outcome.

**Conclusion:** This largest ever case series of COVID-19 patients identified risk factors for COVID-19 diagnosis, hospitalization and adverse outcome. The findings could provide insight for the priorities the need to be set, especially by LMICs, to tackle the pandemic.

## Introduction

As the global pandemic of the Corona Virus Disease 2019 (COVID-19), a disease caused by the Severe Acute Respiratory Syndrome Coronavirus 2 (SARS-CoV-2), is evolving, it is important to understand the pathophysiology and the mechanisms of disease progression. The rapid transmission of the disease and the increased pressure across healthcare systems have led to emergency measures resulting in substantial social and economic disruption. As of May 30, almost 5.9 million people globally have been diagnosed with COVID-19 and approximately 367,255 deaths have been reported. The disease has a wide range of clinical presentations, from mild symptoms resembling the common flu to severe, life-threatening manifestations such as Adult Respiratory Distress Syndrome (ARDS), thrombotic complications and neurological symptoms.^1–3^ Risk factors for adverse outcomes include age, hypertension, diabetes, cardiovascular disease and respiratory disease.^4^

The pandemic represents a big challenge particularly for low- and middle-income countries (LMIC). The cost of epidemiologic surveillance and of infection prevention and control, the sudden flow and the need to enhance the constrained critical care capacity to treat COVID-19 patients and the implementation of non-medical interventions such as social distancing measures are expected to significantly stress the limited financial resources of these countries.^5^ Therefore, understanding the factors associated with COVID-19 susceptibility and adverse prognosis is crucial to guide local authorities towards more efficient allocation of their scarce resources to avoid exceeding the limited capacity of the healthcare system.

In this study, we present evidence and information about patients who were screened for COVID-19 due to suspected viral respiratory infection through the respective surveillance system implemented in Mexico. The objective of this study was to examine the association between individuals’ sociodemographic and clinical characteristics and COVID-19 diagnosis. Additionally, we explored similar associations with two clinical outcomes, hospitalization and adverse outcome, within the COVID-19 diagnosed cohort. This study extends the current literature by presenting novel information and evidence about COVID-19 patients in Mexico using a large and recent cohort of such patients, with potentially important implications at the clinical and policy level.

## Methods

### Study design, setting and participants

We performed a cross-sectional secondary data analysis using a publicly available individual-level dataset which included patients classified as ‘suspected cases of viral respiratory disease’ during point of service at medical facilities in Mexico. The dataset was released by the Mexican Health Ministry and was compiled by the General Bureau of Epidemiology (Dirección General de Epidemiología, DGE) through the System of Epidemiological Surveillance of Viral Respiratory Diseases.^6,7^ The latter comprises of 475 Monitoring Units of Viral Respiratory Disease (Unidades Monitoras de Enfermedad Respiratoria Viral, USMER) spread across the country and covering all institutions affiliated to the Health Ministry collectively denoted as the Health Sector. Additional data were provided by healthcare units that did not belong to USMER but had been adapted to screen suspected COVID-19 cases. As specified by the official guidelines issued by the DGE, the entries in the dataset only correspond to data obtained from the epidemiological study of “suspected case of viral respiratory disease” at the time it was identified at medical units of the Health Sector.

This dataset is continuously updated, and we used the version released on May 31, 2020, which included 274,997 patients.

No ethics approval was sought for this study since it involves analysis of an anonymized dataset of patients that is publicly available and accessible to anyone through the Mexican Health Ministry.

### Data sources

Upon admission, patients were screened by healthcare professionals who were expected to verify that the subjects show specific symptoms documented as inclusion criteria for the dataset. Additionally, they recorded data about the medical history on a specific DGE form. After the case was evaluated and confirmed at the district, state and national level surveillance system, it was added to the dataset. Both USMER and Non-USMER units had to fill the same forms which were sent to an online platform (SISVER platform). According to the DGE guidance, USMER and non-USMER units should perform diagnostic testing for COVID-19 (RT-PCR) in all cases with serious symptoms. For cases with mild symptoms (classified as ambulatory cases), USMER units were expected to perform COVID-19 diagnostic testing on 10% of these cases whereas non-USMER units would test cases depending on their resource capacity.

Laboratory testing to confirm SARS-CoV-2 infection was performed according to WHO interim guidance.^8^ Combined nasopharyngeal and oropharyngeal swabs were obtained and placed in a container. For mechanically-ventilated patients, bronchoalveolar lavage was obtained. In case of death, lung biopsies were obtained during autopsy, from an area visibly affected by disease. The samples were sent to the nearest Laboratory of Respiratory Virus (InDRE) for testing with RTPCR.

### Variables

The dataset included information on COVID-19 testing results, categorized as positive, negative and pending, and individual-level data on sociodemographics, specifically age, gender and nationality (Mexican or not). The type of facility where the patient was diagnosed was reported, as well as information on whether the healthcare unit was part of the USMER network or not. Comorbidities reported were diabetes, chronic obstructive lung disease (COPD), asthma, immunosuppression, hypertension, cardiovascular disease, obesity, chronic renal disease and other comorbidities. Smoking was also recorded, with participants classified as smokers (who were considered current smokers) or non-smokers. No data on former smokers was available. For patients with laboratory-confirmed COVID-19 diagnosis, the dataset included additional information related to clinical endpoints, namely whether the patient was admitted into an intensive care unit, was mechanically ventilated, or died.

### Outcomes and analysis

The first outcome variable of interest in the study was whether a patient was diagnosed with COVID-19 or not, defined as a dichotomous indicator. We thus excluded 36,803 patients (13.4%) with pending results, resulting in a final sample of 238,194 patients. We also explored two outcomes within the subgroup of patients with COVID-19 diagnosis, hospitalization and adverse outcome. Both outcomes were dichotomous. Adverse outcome was defined as the primary composite end-point of intensive care unit admission, mechanical ventilation or death.

We included individual-level sociodemographic and clinical characteristics, and facility information based on data availability. Sociodemographic patient-level information included age, gender, and Mexican nationality (or not). Clinical information included number of comorbidities, whether the patient had a particular clinical condition or not, namely asthma, chronic obstructive pulmonary disease (COPD), diabetes, obesity, hypertension, immunosuppression, cardiovascular condition, and chronic renal disease. We also included information on whether the patient was a current smoker and whether there was previous contact with someone who was diagnosed with COVID-19. Facility specific information included the type of facility by ownership and whether the medical unit is a monitoring unit for respiratory diseases (USMER). USMER consists of medical facilities which monitor the incidence of infectious respiratory diseases as part of the government Epidemiological Surveillance system.

We initially conducted descriptive analyses of all patients to characterize the overall study population. We then explored and compared all patients stratified by whether they were diagnosed with COVID-19 or not. Subsequently, laboratory-confirmed COVID-19 cases were stratified by the two outcomes of interest (hospitalization and adverse outcome) using bivariate analyses. We tested for statistical differences in the stratified analyses using Pearson’s chi-square for categorical variables and the non-parametric Mann Whitney U test on the age variable (as numeric).

We then estimated the association between the three outcomes of interest and all the independent variables using two multivariate logistic regressions per outcome (six in total). Both models included sociodemographic and facility-specific information. The first model also included clinical comorbidities as dichotomous indicators, while the second included the number of clinical diagnoses (comorbidities) only, due to multicollinearity as the later was derived from the clinical diagnoses. We also included area-of-residence fixed-effects to control for unobserved regional variations. Finally, standard errors were clustered at the residence level. All analyses were conducted using Stata (version 16.1; StataCorp, College Station, TX).

## Results

Our final analytic sample included 236,439 patients suspected of having viral respiratory disease, after excluding 1,755 patients (0.7%) due to missing variables. The majority were 18 to 44 years of age, Mexicans, and around 40% of those had one or more comorbidities, while 9.1% were current smokers (**Table 1**). The most prevalent clinical conditions included hypertension, diabetes, and obesity. About 37% of the patients used a USMER facility and more than half used hospitals of the Ministry of Health (Secretaria de Salubridad y Asistencia – SSA).

**Table 1.**
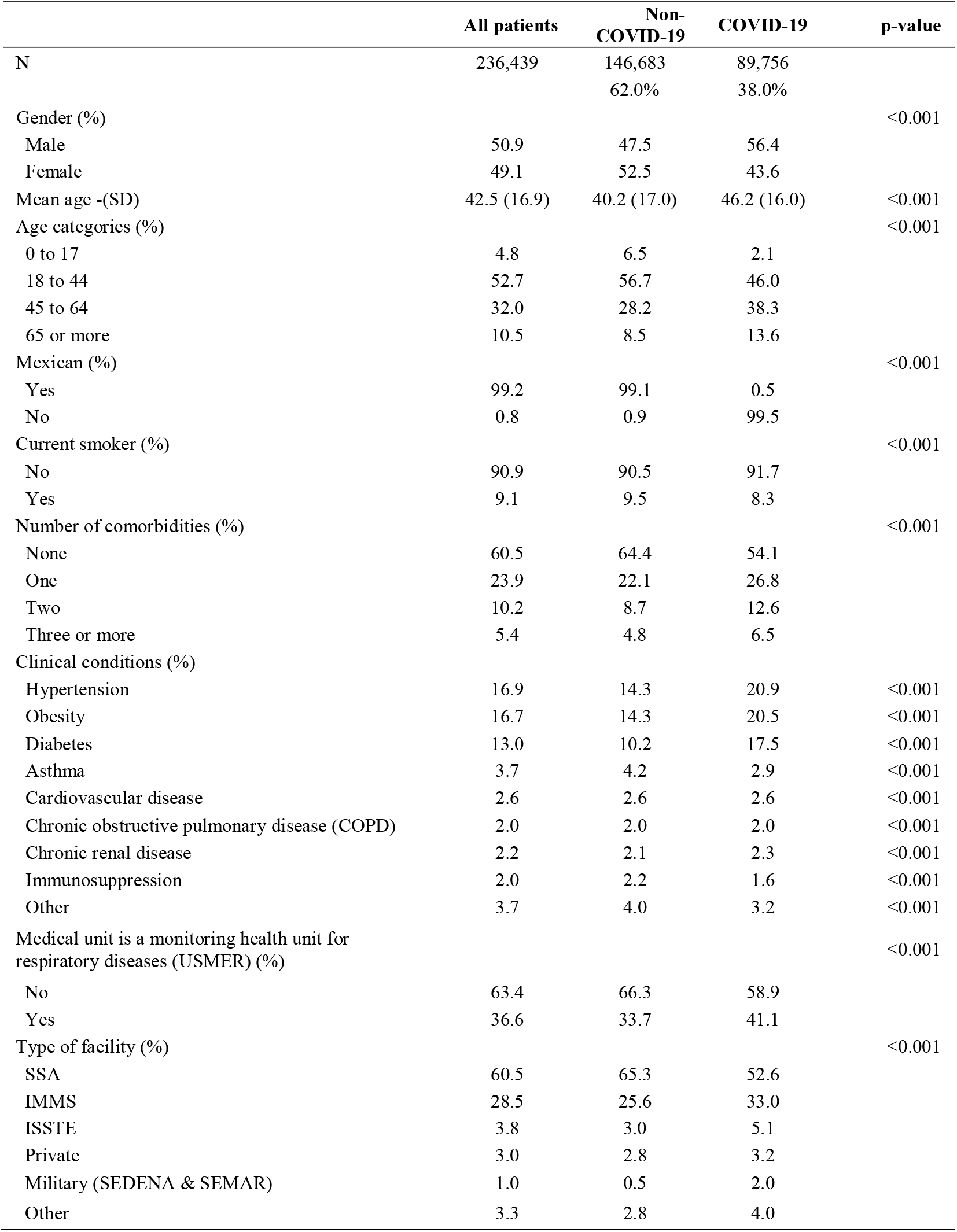
Descriptive analysis on all patients with suspected viral respiratory disease and bivariate analysis stratified by positive or negative COVID-19 diagnosis.

|  | All patients | Non-COVID-19 | COVID-19 | p-value |
| --- | --- | --- | --- | --- |
| N | 236,439 | 146,683<br>62.0% | 89,756<br>38.0% |  |
| Gender (%) |  |  |  | <0.001 |
| Male | 50.9 | 47.5 | 56.4 |  |
| Female | 49.1 | 52.5 | 43.6 |  |
| Mean age -(SD) | 42.5 (16.9) | 40.2 (17.0) | 46.2 (16.0) | <0.001 |
| Age categories (%) |  |  |  | <0.001 |
| 0 to 17 | 4.8 | 6.5 | 2.1 |  |
| 18 to 44 | 52.7 | 56.7 | 46.0 |  |
| 45 to 64 | 32.0 | 28.2 | 38.3 |  |
| 65 or more | 10.5 | 8.5 | 13.6 |  |
| Mexican (%) |  |  |  | <0.001 |
| Yes | 99.2 | 99.1 | 0.5 |  |
| No | 0.8 | 0.9 | 99.5 |  |
| Current smoker (%) |  |  |  | <0.001 |
| No | 90.9 | 90.5 | 91.7 |  |
| Yes | 9.1 | 9.5 | 8.3 |  |
| Number of comorbidities (%) |  |  |  | <0.001 |
| None | 60.5 | 64.4 | 54.1 |  |
| One | 23.9 | 22.1 | 26.8 |  |
| Two | 10.2 | 8.7 | 12.6 |  |
| Three or more | 5.4 | 4.8 | 6.5 |  |
| Clinical conditions (%) |  |  |  |  |
| Hypertension | 16.9 | 14.3 | 20.9 | <0.001 |
| Obesity | 16.7 | 14.3 | 20.5 | <0.001 |
| Diabetes | 13.0 | 10.2 | 17.5 | <0.001 |
| Asthma | 3.7 | 4.2 | 2.9 | <0.001 |
| Cardiovascular disease | 2.6 | 2.6 | 2.6 | <0.001 |
| Chronic obstructive pulmonary disease (COPD) | 2.0 | 2.0 | 2.0 | <0.001 |
| Chronic renal disease | 2.2 | 2.1 | 2.3 | <0.001 |
| Immunosuppression | 2.0 | 2.2 | 1.6 | <0.001 |
| Other | 3.7 | 4.0 | 3.2 | <0.001 |
| Medical unit is a monitoring health unit for<br>respiratory diseases (USMER) (%) |  |  |  | <0.001 |
| No | 63.4 | 66.3 | 58.9 |  |
| Yes | 36.6 | 33.7 | 41.1 |  |
| Type of facility (%) |  |  |  | <0.001 |
| SSA | 60.5 | 65.3 | 52.6 |  |
| IMMS | 28.5 | 25.6 | 33.0 |  |
| ISSTE | 3.8 | 3.0 | 5.1 |  |
| Private | 3.0 | 2.8 | 3.2 |  |
| Military (SEDENA & SEMAR) | 1.0 | 0.5 | 2.0 |  |
| Other | 3.3 | 2.8 | 4.0 |  |

Around one-third (38.0%) of patients were diagnosed with COVID-19 (Table 1). These patients had higher shares of males (56.4% versus 47.5% for non-COVID-19 patients, p< 0.001), and were 6 years older on average (p< 0.001). COVID-19 patients had also disproportionately higher shares of one or more comorbidities (p< 0.001), and chronic conditions particularly related to diabetes, hypertension, and obesity (p< 0.001 for all). We also observed greater shares for USMER related COVID-19 cases at the facility level (p< 0.001 for both).

**Table 2** indicates the results of the two regressions on COVID-19 diagnosis for all patients in the data. Across both models, females and younger patients (0 to 17) were significantly less likely to be diagnosed with COVID-19 compared to males and to their 18 to 44 counterparts (p< 0.001for all). In addition, current smokers were approximately 23% less likely to be diagnosed with COVID-19 compared to non-smokers (p< 0.001 in both models). In contrast, older patients (45 years of age or older) and those with one or more comorbidities were more likely to be diagnosed with COVID-19 compared to those aged 18 to 44 and those without comorbidities respectively. Diabetes and obesity were particularly associated with COVID-19 diagnosis compared to patients without such conditions (p< 0.001 for all).

**Table 2.** Multivariate logistic regression analyses on the factors associated with COVID-19 diagnosis across all patients. Two regression models were examined, one with each comorbidity introduced separately as independent variable and one with number of comorbidities used as independent variable.

|  | OR | 95%CI | p-value | OR | 95%CI | p-value |
| --- | --- | --- | --- | --- | --- | --- |
| Gender (Ref: Male) |  |  |  |  |  |  |
| Female | 0.70 | 0.67-0.72 | <0.001 | 0.69 | 0.67-0.72 | <0.001 |
| Age (Ref: 18 to 44) |  |  |  |  |  |  |
| 65 or more | 1.73 | 1.60-1.88 | <0.001 | 1.66 | 1.52-1.81 | <0.001 |
| 45 to 64 | 1.46 | 1.37-1.55 | <0.001 | 1.48 | 1.40-1.57 | <0.001 |
| 0 to 17 | 0.47 | 0.40-0.56 | <0.001 | 0.44 | 0.36-0.53 | <0.001 |
| Mexican (Ref: Yes) |  |  |  |  |  |  |
| No | 0.58 | 0.50-0.68 | <0.001 | 0.56 | 0.47-0.65 | <0.001 |
| Smoker (Ref: No) |  |  |  |  |  |  |
| Yes | 0.77 | 0.74-0.80 | <0.001 | 0.77 | 0.73-0.80 | <0.001 |
| Obese (Ref: No) |  |  |  |  |  |  |
| Yes | 1.38 | 1.33-1.42 | <0.001 | - | - | - |
| Diabetes (Ref: No) |  |  |  |  |  |  |
| Yes | 1.36 | 1.31-1.41 | <0.001 | - | - | - |
| Hypertension (Ref: No) |  |  |  |  |  |  |
| Yes | 1.04 | 1.01-1.08 | 0.006 |  |  |  |
| Immunosuppression (Ref: No) |  |  |  |  |  |  |
| Yes | 0.70 | 0.63-0.78 | <0.001 | - | - | - |
| Cardiovascular disease (Ref: No) |  |  |  |  |  |  |
| Yes | 0.77 | 0.72-0.82 | <0.001 | - | - | - |
| Asthma (Ref: No) |  |  |  |  |  |  |
| Yes | 0.72 | 0.66-0.78 | <0.001 | - | - | - |
| COPD (Ref: No) |  |  |  |  |  |  |
| Yes | 0.73 | 0.63-0.85 | <0.001 | - | - | - |
| Chronic renal disease (Ref: No) |  |  |  |  |  |  |
| Yes | 0.81 | 0.69-0.95 | <0.001 | - | - | - |
| Number of comorbidities (Ref: None) |  |  |  |  |  |  |
| One | - | - | - | 1.20 | 1.14-1.27 | <0.001 |
| Two | - | - | - | 1.26 | 1.17-1.34 | <0.001 |
| Three or more | - | - | - | 1.17 | 1.04-1.30 | 0.007 |
| Medical unit is a monitoring health unit for respiratory diseases (USMER) (Ref: No) |  |  |  |  |  |  |
| Yes | 1.24 | 1.10-1.40 | 0.001 | 1.24 | 1.10-1.39 | <0.001 |
| Type of facility (Ref: SSA) |  |  |  |  |  |  |
| IMMS | 1.48 | 1.24-1.75 | <0.001 | 1.42 | 1.20-1.68 | <0.001 |
| ISSTE | 1.51 | 1.12-2.03 | 0.007 | 1.48 | 1.09-2.0 | 0.011 |
| Private | 1.38 | 1.17-1.64 | <0.001 | 1.34 | 1.13-1.58 | 0.001 |
| Military (SEDENA & SEMAR) | 5.06 | 2.97-8.63 | <0.001 | 4.95 | 2.87-8.54 | <0.001 |
| Other | 1.24 | 0.95-1.63 | 0.117 | 1.25 | 0.96-1.63 | 0.099 |

**Notes:** Both analyses control for area-of-residence fixed-effects; OR: Odds ratio; CI: Confidence Intervals.

Among the 89,756 patients who were diagnosed with COVID-19, about 35% were hospitalized and 13% had high clinical severity (**Table 3**). Across both subgroups, hospitalization and adverse outcome were more frequent in males and older patients, with a mean age difference of more than 12 years. Patients with one or more comorbidities, particularly those with hypertension, obesity, diabetes, and COPD were also more prevalent in both the hospitalized and the adverse outcome groups.

**Table 3.** Descriptive bivariate analysis among patients with COVID-19 diagnosis stratified by hospitalization and adverse outcome.

|  | All<br>COVID-19<br>diagnosed | Hospitalized |  | p-value | Adverse outcome |  | p-value |
| --- | --- | --- | --- | --- | --- | --- | --- |
|  |  | No | Yes |  | No | Yes |  |
| N | 89,756 | 58,485<br>(65.2%) | 31,271<br>(34.8%) |  | 78,050<br>(87.0%) | 11,706<br>(13.0%) |  |
| Gender (%) |  |  |  | <0.001 |  |  | <0.001 |
| Male | 56.4 | 52.4 | 63.9 |  | 54.8 | 67.0 |  |
| Female | 43.6 | 47.6 | 36.1 |  | 45.2 | 33.0 |  |
| Mean age | 46.2 (16.0) | 41.8 (14.4) | 54.5 (15.5) | <0.001 | 44.3 (15.3) | 58.9 (14.8) | <0.001 |
| Age categories (%) |  |  |  | <0.001 |  |  | <0.001 |
| 0 to 17 | 2.1 | 2.6 | 1.1 |  | 2.3 | 0.8 |  |
| 18 to 44 | 46.0 | 57.8 | 24.0 |  | 50.7 | 17.7 |  |
| 45 to 64 | 38.3 | 32.9 | 48.2 |  | 36.8 | 48.2 |  |
| 65 or more | 13.6 | 6.7 | 26.7 |  | 10.2 | 36.3 |  |
| Mexican |  |  |  | <0.001 |  |  | 0.009 |
| Yes | 99.5 | 99.5 | 99.7 |  | 99.5 | 99.7 |  |
| No | 0.5 | 0.5 | 0.3 |  | 0.5 | 0.3 |  |
| Current smoker (%) |  |  |  | 0.002 |  |  | 0.002 |
| No | 91.7 | 92 | 91.3 |  | 91.9 | 90.7 |  |
| Yes | 8.3 | 8.0 | 8.7 |  | 8.1 | 9.3 |  |
| Clinical conditions (%) |  |  |  |  |  |  |  |
| Hypertension | 20.9 | 14.3 | 33.3 | <0.001 | 18.0 | 40.5 | <0.001 |
| Obesity | 20.5 | 18.6 | 24.1 | <0.001 | 19.5 | 27.0 | <0.001 |
| Diabetes | 17.5 | 10.6 | 30.4 | <0.001 | 14.6 | 36.7 | <0.001 |
| Asthma | 2.9 | 3.2 | 2.4 | <0.001 | 3.0 | 2.3 | <0.001 |
| Cardiovascular disease | 2.6 | 1.7 | 4.2 | <0.001 | 2.2 | 5.5 | <0.001 |
| Chronic renal disease | 2.3 | 1.1 | 4.7 | <0.001 | 1.7 | 6.5 | <0.001 |
| Chronic obstructive pulmonary disease (COPD) | 2.0 | 1.0 | 3.8 | <0.001 | 1.6 | 4.9 | <0.001 |
| Immunosuppression | 1.6 | 1.0 | 2.6 | <0.001 | 1.4 | 2.9 | <0.001 |
| Other | 3.2 | 2.6 | 4.3 | <0.001 | 3.0 | 4.7 | <0.001 |
| Number of comorbidities (%) |  |  |  | <0.001 |  |  | <0.001 |
| None | 54.1 | 63.1 | 37.2 |  | 57.8 | 28.9 |  |
| One | 26.8 | 24.4 | 31.1 |  | 26.0 | 32.0 |  |
| Two | 12.6 | 8.9 | 19.7 |  | 11.1 | 23.1 |  |
| Three or more | 6.5 | 3.6 | 12.0 |  | 5.1 | 16.0 |  |
| Medical unit is a monitoring health unit for respiratory diseases (USMER) (%) |  |  |  | <0.001 |  |  | <0.001 |
| No | 58.9 | 68.6 | 40.6 |  | 61.8 | 39.4 |  |
| Yes | 41.1 | 31.4 | 59.4 |  | 38.2 | 60.6 |  |
| Type of facility (%) |  |  |  | <0.001 |  |  | <0.001 |
| SSA | 52.6 | 60.0 | 38.7 |  | 53.6 | 45.6 |  |
| IMMS | 33.0 | 28.4 | 41.7 |  | 32.7 | 35.4 |  |
| ISSTE | 5.1 | 2.7 | 9.5 |  | 4.5 | 9.1 |  |
| Private | 3.2 | 3.3 | 3.1 |  | 3.2 | 3.4 |  |
| Military | 2.0 | 2.2 | 1.6 |  | 2.1 | 1.4 |  |
| Other | 4.0 | 3.3 | 5.5 |  | 3.9 | 5.1 |  |

**Table 4** indicates the results of the two regressions for hospitalization and adverse outcome, respectively (Table 4). Across both models, males and older patients were significantly more likely to be hospitalized and to experience adverse outcome compared to females and to their 18 to 44 old counterparts (p< 0.001 for all). Those 0 to 17 years of age were also less likely to experience adverse outcome compared to the 18 to 44 years age group. In addition, patients with chronic renal disease, diabetes, immunosuppression, COPD, obesity, and hypertension were up to 121% (adjusted OR: 2.21, 95% CI: 1.91–2.55, for those with chronic renal disease) more likely to experience hospitalization and more severe composite endpoints compared to those without such conditions (p< 0.001 for all). Similarly, having one or more comorbidities increased the likelihood of these outcomes, as expected (**Table 5**). Finally, we did not observe any significant differences among current smokers compared to non-smokers across both outcomes.

**Table 4.** Multivariate logistic regression analyses of factors associated with hospitalization and adverse outcome among patients with COVID-19 diagnosis.

|  | Hospitalized |  |  | Adverse outcome |  |  |
| --- | --- | --- | --- | --- | --- | --- |
|  | OR | 95%CI | p-value | OR | 95%CI | p-value |
| Gender (Ref: Male) |  |  |  |  |  |  |
| Female | 0.57 | 0.53-0.61 | <0.001 | 0.58 | 0.53-0.64 | <0.001 |
| Age (Ref: 18 to 44) |  |  |  |  |  |  |
| 65 or more | 7.24 | 6.22-8.42 | <0.001 | 8.30 | 7.12-9.67 | <0.001 |
| 45 to 64 | 2.93 | 2.76-3.12 | <0.001 | 3.57 | 3.17-3.79 | <0.001 |
| 0 to 17 | 1.73 | 1.39-2.14 | <0.001 | 1.52 | 1.16-2.00 | 0.003 |
| Mexican (Ref: Yes) |  |  |  |  |  |  |
| No | 0.86 | 0.65-1.12 | 0.263 | 0.73 | 0.52-1.01 | 0.06 |
| Smoker (Ref: No) |  |  |  |  |  |  |
| Yes | 0.94 | 0.85-1.04 | 0.219 | 1.02 | 0.88-1.18 | 0.827 |
| Chronic renal disease (Ref: No) |  |  |  |  |  |  |
| Yes | 2.21 | 1.91-2.55 | <0.001 | 1.96 | 1.69-2.26 | <0.001 |
| Diabetes (Ref: No) |  |  |  |  |  |  |
| Yes | 1.99 | 1.85-2.14 | <0.001 | 1.64 | 1.55-1.73 | <0.001 |
| Immunosuppression (Ref: No) |  |  |  |  |  |  |
| Yes | 1.85 | 1.59-2.15 | <0.001 | 1.41 | 1.22-1.64 | 0.006 |
| COPD (Ref: No) |  |  |  |  |  |  |
| Yes | 1.44 | 1.26-1.65 | <0.001 | 1.33 | 1.20-1.49 | <0.001 |
| Obese (Ref: No) |  |  |  |  |  |  |
| Yes | 1.40 | 1.29-1.51 | <0.001 | 1.51 | 1.39-1.64 | <0.001 |
| Hypertension (Ref: No) |  |  |  |  |  |  |
| Yes | 1.25 | 1.16-1.34 | <0.001 | 1.28 | 1.20-1.35 | <0.001 |
| Cardiovascular disease (Ref: No) |  |  |  |  |  |  |
| Yes | 1.09 | 0.96-1.24 | 0.185 | 1.09 | 1.01-1.17 | 0.031 |
| Asthma (Ref: No) |  |  |  |  |  |  |
| Yes | 0.73 | 0.65-0.81 | <0.001 | 0.82 | 0.71-0.94 | 0.003 |
| Medical unit is a monitoring health unit for respiratory diseases (USMER) (Ref: No) |  |  |  |  |  |  |
| Yes | 3.57 | 2.85-4.48 | <0.001 | 2.32 | 1.88-2.85 | <0.001 |
| Type of facility (Ref: SSA) |  |  |  |  |  |  |
| IMMS | 2.17 | 1.31-3.58 | 0.003 | 0.99 | 0.76-1.29 | 0.944 |
| ISSTE | 3.32 | 1.29-8.52 | 0.013 | 1.23 | 0.79-1.93 | 0.357 |
| Private | 1.82 | 0.89-3.71 | 0.099 | 1.45 | 1.07-1.96 | 0.017 |
| Military (SEDENA & SEMAR) | 2.33 | 1.41-3.83 | 0.001 | 1.38 | 0.87-2.18 | 0.177 |
| Other | 1.91 | 1.20-3.03 | 0.006 | 1.01 | 0.74-1.38 | 0.937 |

**Notes:** Both analyses control for area-of-residence fixed-effects; OR: Odds ratio; CI: Confidence Intervals.

**Table 5.** Multivariate logistic regression analyses of factors associated with hospitalization and adverse outcome among patients with COVID-19 diagnosis, in which number of comorbidities (instead of each comorbidity) was used as independent variable.

|  | Hospitalized |  |  | Adverse outcome |  |  |
| --- | --- | --- | --- | --- | --- | --- |
|  | OR | 95%CI | p-value | OR | 95%CI | p-value |
| Number of comorbidities (Ref: None) |  |  |  |  |  |  |
| One | 1.72 | 1.57-1.89 | <0.001 | 1.79 | 1.69-1.90 | <0.001 |
| Two | 2.37 | 2.09-2.67 | <0.001 | 2.36 | 2.15-2.59 | <0.001 |
| Three or more | 3.29 | 2.83-3.81 | <0.001 | 3.35 | 2.96-3.78 | <0.001 |

**Notes:** Both models control for all other covariates in the main models in Table 4; OR: Odds ratio; CI: Confidence Intervals

## Discussion

To the best of our knowledge, this study presents a case series with the highest number of laboratory-confirmed COVID-19 patients, and the first of this size from a LMIC. The Chinese Centers for Disease Control (CDC) recently presented data from 44,672 confirmed cases, however information about comorbidities was available for only 20,812 patients [9]. Another study in the UK examined risk factors for critical care and mortality in hospital among 20,133 hospitalized COVID-19 patients [10]. In the US, patient characteristics, comorbidities and outcomes were presented among 5,700 patients hospitalized for COVID-19 in New York City area [11]. Our study adds to the current evidence by presenting information from laboratory-confirmed cases in a LMIC using a large publicly available dataset.

In agreement with previous studies [1, 10–15], age was a strong risk factor for hospitalization and adverse outcome among COVID-19 patients. It was recognized early during the pandemic that the elderly had higher rates of hospitalization and infection fatality ratios compared to younger people [16]. Recently, the US CDC reported that the best estimate for the COVID-19 symptomatic case fatality ratio was 0.4% for the whole population but ranged from 0.05% for those aged 0–49 years to 1.3% for those ≥ 65 years, a 26-fold difference [17]. Hospitalization rates were estimated to be approximately almost 7-fold higher for patients aged ≥ 65 years compared to those aged 18 to 44 years in our study. Thus, targeted interventions tailored at the higher needs of older people are clearly to protect them from SARS-CoV-2 infection and to reduce the COVID-19 burden of disease and death rates.

Our results indicate that cardiovascular and metabolic conditions were the most common comorbidities identified among confirmed COVID-19 patients. Hypertension, obesity and diabetes were not only common comorbidities but also independent correlates of hospitalization ≥ and adverse outcome. These findings are in-line with case series from China, the US and Europe [1, 9–14]. These conditions are very common worldwide and in Mexico. Approximately 1.4 billion adults are estimated to suffer from hypertension globally, with the prevalence being higher in LMICs [18]. In Mexico the prevalence of hypertension was 25.5% in 2016 [19]. Obesity is also a major healthcare issue in Mexico. In a random sample of 2,511 adults, 38.3% of Mexicans were overweight and almost 25% were obese [20]. The Organization for Economic Cooperation and Development (OECD) reports that Mexico is the one of the countries with the highest rates of obesity in the population [21]. Obesity is a risk factor for diabetes, and Mexico had an estimated 10.4% prevalence of diabetes in 2016 with a continuously increasing trend [22, 23]. The latter, in combination of increased prevalence of these conditions among COVID-19 patients suggests that such patients represent another population subgroup where targeted interventions and guidance are needed to prevent SARS-CoV-2 transmission.

In contrast, while cardiovascular disease and COPD were risk factors for hospitalization and adverse outcome, only a small proportion of patients suffered from these comorbidities. A case series of 1,590 patients from China reported a similarly low prevalence of these comorbidities among Chine patients [12]. The COPD prevalence in Mexico City was 3.4% in a study defining airflow obstruction as FEV1/FEV_6_ below the 5th percentile or Lower Limit of Normal [24], but it has been reported that COPD is highly underdiagnosed in Mexico and in other countries, mainly because of lack of spirometry evaluation [25]. In a 2009 study, ischemic heart disease and stroke prevalence in Mexico City ranged from 0.4% to 5.4% and from 0.4% to 3.5%, respectively, depending on age [26]. In addition, other risk factors for adverse outcomes were immunosuppression and chronic renal disease. Our findings are supported by a recent systematic review and meta-analysis which found a higher risk for adverse COVID-19 outcomes among patients with chronic renal disease [27].

Furthermore, having more than one comorbidity was strongly associated with hospitalization and adverse outcome. This is not unexpected considering that multiple comorbidities contribute to disease complexity and such patients are more susceptible and vulnerable to adverse events. Approximately 1 in 5 patients with laboratory-diagnosed COVID-19 had more than 1 comorbidity, and they had approximately 3-fold higher risk for hospitalization and adverse outcome. Therefore, it is necessary to prioritize the assessment of these patients, offering early diagnosis and proper hospital care, while primary preventive measures to reduce disease transmission to these patients are warranted.

Notably, smoking was not associated with a higher risk for adverse outcomes and hospitalization. Smokers were also less likely to be diagnosed with COVID-19 compared to nonsmokers. The latter is in agreement with a recent observational population study from Israel [28]. Some studies have found that smokers are under-represented among COVID-19 patients and presented a hypothesis that nicotine may exert protective effects [29–31], while others found that nicotine and smoking cause ACE2 up-regulation which may facilitate viral invasion and cell entry [32, 33]. It is still not clear whether nicotine exerts any positive effect or not. However, there is no doubt that smoking cannot be used as a protective measure and smoking cessation should be encouraged during the COVID-19 pandemic [30]. The potential value of nicotine or other nicotinic agonists in COVID-19 is expected to be determined through clinical trials.

It has been established that SARS-CoV-2 uses angiotensin-converting enzyme 2 (ACE2) as a receptor for cell entry and viral replication [34]. While this would imply that ACE2 upregulation would be associated with COVID-19 severity and adverse outcome, there is evidence that the opposite is the case. Risk factors for adverse outcomes identified in this and other studies, such as age, male gender, endocrine and cardiovascular diseases, are associated with lower levels of ACE2. [35–38]. Therefore, it has been hypothesized that ACE2 deficiency may in fact be detrimental for COVID-19 [39]. Additionally, severe COVID-19 represents a hyper-inflammatory response with patients developing cytokine storm and exhibiting ineffective regulation of the immune response [40]. Risk factors identified in this study also represent inflammatory conditions [41–44]. Thus, there is a relevant pathophysiological basis explaining the association between hospitalized and adverse COVID-19 outcomes and the comorbidities identified in our analysis.

This study is not without limitations. First, given the nature and the availability of the data, we were not able to use more detailed clinical and laboratory information for the patients. For example, immunodeficiency may include a vast array of different disease conditions; however, no specific information was provided in the dataset. Second, we were not able to include facility and regional specific information, due to the lack of such information in the dataset. However, we believe that we sufficiently addressed the heterogeneity between hospitals and regions using the appropriate variables in our analyses, given the research question of interest. Finally, some of the patients may not have recovered by the time the dataset was released and thus the outcome is unknown, while it is also possible that some outpatients may experience disease progression and will thus require hospitalization in the future.

In conclusion, this large retrospective case series from Mexico, the largest ever presented for COVID-19, identified risk factors for laboratory-confirmed COVID-19 diagnosis as well as for hospitalization and adverse outcome among COVID-19 patients. These findings could provide valuable insight for Mexico and other LMICs in setting priorities, properly allocating healthcare resources and establishing disease transmission preventive strategies in order to protect vulnerable groups, particularly the elderly and people with comorbidities.

## Data Availability

The dataset is publicly available and accessible to anyone through the Mexican Health Ministry.

